# Creation of synthetic contrast-enhanced computed tomography images using deep neural networks to screen for renal cell carcinoma

**DOI:** 10.1101/2022.01.12.22269120

**Authors:** Naoto Sassa, Yoshitaka Kameya, Tomoichi Takahashi, Yoshihisa Matsukawa, Tsuyoshi Majima, Katsuhisa Tsuruta, Ikuo Kobayashi, Keishi Kajikawa, Hideji Kawanishi, Haruka Kurosu, Sho Yamagiwa, Masaya Takahashi, Kazuhiro Hotta, Keiichi Yamada, Tokunori Yamamoto

## Abstract

**Objectives:** To elucidate if synthetic contrast enhanced computed tomography (CECT) images created from plain CT images using deep neural networks (DNN) could be used for screening, clinical diagnosis, and postoperative follow-up of small-diameter renal tumors by comparing the concordance rate between real and synthetic CECT images and the diagnoses according to 10 urologists.

**Methods:** This retrospective, multicenter study included 155 patients (artificial intelligence training cohort [n=99], validation cohort [n=56]) who underwent surgery for small-diameter (≤40 mm) renal tumors, with the pathological diagnosis of renal cell carcinoma, during 2010–2020. Preoperatively, dynamic plain CT and CECT images were obtained. We created a learned DNN using pix2pix. We examined the quality of the synthetic CECT images created using this DNN and compared them with real CECT images using the zero-mean normalized cross-correlation parameter. We assessed concordance rates between real and synthetic images and diagnoses according to 10 urologists by creating a receiver operating characteristic curve and calculating the area under the curve (AUC).

**Results:** The synthetic CECT images were highly concordant with the real CECT images, regardless of the existence or morphology of the renal tumor. Regarding the concordance rate, a greater AUC was obtained with synthetic CECT (AUC=0.892) than with only CT (AUC=0.720; p<0.001).

**Conclusions:** This study is the first to use DNN to create a high-quality synthetic CECT image that was highly concordant with a real CECT image. Synthetic CECT images could be used for urological diagnoses and clinical screening.

## Introduction

Recently, the number of accidentally discovered small-diameter renal tumors has increased^1^ More than 50% of kidney tumors are asymptomatic or discovered while screening for other illnesses.^2,3^ It is necessary to perform both plain computed tomography (CT), which does not use a contrast medium, and contrast-enhanced CT (CECT), which uses a contrast medium, to diagnose renal cell carcinoma (RCC).^4^ The methods of CECT were determined by different clinical indications according to the renal protocol, balancing diagnostic accuracy and radiation exposure.^5^ CECT shows the presence or absence of blood flow in the tumor by comparing the Hounsfield units (*HU)* before and after the injection of the contrast medium; an enhancement of the contrast effect by ≥15 HU when compared with plain CT indicates the presence of a kidney tumor.^6^ Additionally, CECT angiography is useful for visualizing the location of blood vessels before surgery.^7^ However, the use of a contrast medium is contraindicated in patients with contrast medium-related allergies and moderate renal dysfunction.^8^ Moreover, RCC also occurs in younger patients, and thus, these patients are subjected to frequent medical exposure to CT during screening and follow-up following radical surgery. Imaging methods aimed at reducing medical exposure have been attempted previously.^9^ Magnetic resonance imaging (MRI) is recommended to reduce the risk of secondary carcinogenesis owing to medical exposure.^10-12^ MRI is useful for determining the presence or absence of tumor thrombus in inferior vena cava in patients with RCC; however, its resolution is inferior to that of CECT. Imaging modalities with reduced exposure doses and better image detection capabilities have not yet been developed.^13^ Additionally, the European Association of Urology (EAU) guidelines recommend the development of a postoperative CT schedule according to the risk and frequency of RCC recurrence-based tumor staging to reduce medical exposure.^14^

The progress in image composition technology has been remarkable. There have been many reports in the medical field on improving diagnostic imaging assistance using artificial intelligence (AI). AI is used to distinguish between benign and malignant renal tumors.^15-18^ Some studies have sought to determine the grade and type of malignant and nuclear atypia of RCC.^19,20^ However, all studies utilizing AI have used previously obtained CECT images and not image composition technology. Furthermore, while previous studies have also reported CT image generation by image-to-image translation using deep neural networks (DNNs),^21^ there have been no reports on synthetic CECT images created for the purpose of reducing medical exposure and avoiding the use of a contrast medium. In this study, we first created a DNN based on plain CT images. We subsequently aimed to evaluate whether a synthetic CECT image created using the DNN could be used for clinical diagnosis by comparing the concordance rate between real and synthetic CECT images and the diagnoses made by 10 urologists.

## Material and Methods

This study was approved by the appropriate ethical committees, and verbal informed consent was obtained from each participant. One-hundred fifty-five patients who underwent surgery for small-diameter (≤ 40 mm) renal tumors, with a pathological diagnosis, at Aichi Medical University and Nagoya University between 2010 and 2020 were included. Preoperatively, dynamic plain CT and CECT images were obtained from all patients. Except for one patient whose bilateral kidneys were affected, CT image analysis of each patient revealed a small renal tumor in only one kidney. Patient information, including patient age, sex, tumor laterality, tumor size, tumor location, and R.E.N.AL. nephrometry score, was acquired from the medical records.^22^ In all cases, the diagnosis of renal cancer by CT was determined by the presence or absence of tumor blood flow, tumor morphology, and blood flow pattern visualized using a contrast agent. The pathological diagnosis included pathological malignant or benign tumor, histological type, pathological T stage, and nuclear grade according to the WHO 2016 classification and Fuhrman nuclear grade. All plain CT and CECT (arterial, venous, and urinary excretion phases) images of the chest to the lower abdominal region of 155 patients were obtained in the digital imaging and communications in medicine (DICOM) format. In total, 155 patients with 309 kidneys (one patient had only one kidney with a renal tumor) were divided into two cohorts: the AI training cohort and the evaluation cohort (Figure 1). We compared the concordance rate between synthetic CECT images created using the DNN learned using the AI training cohort and real CECT images.

**Figure 1.**
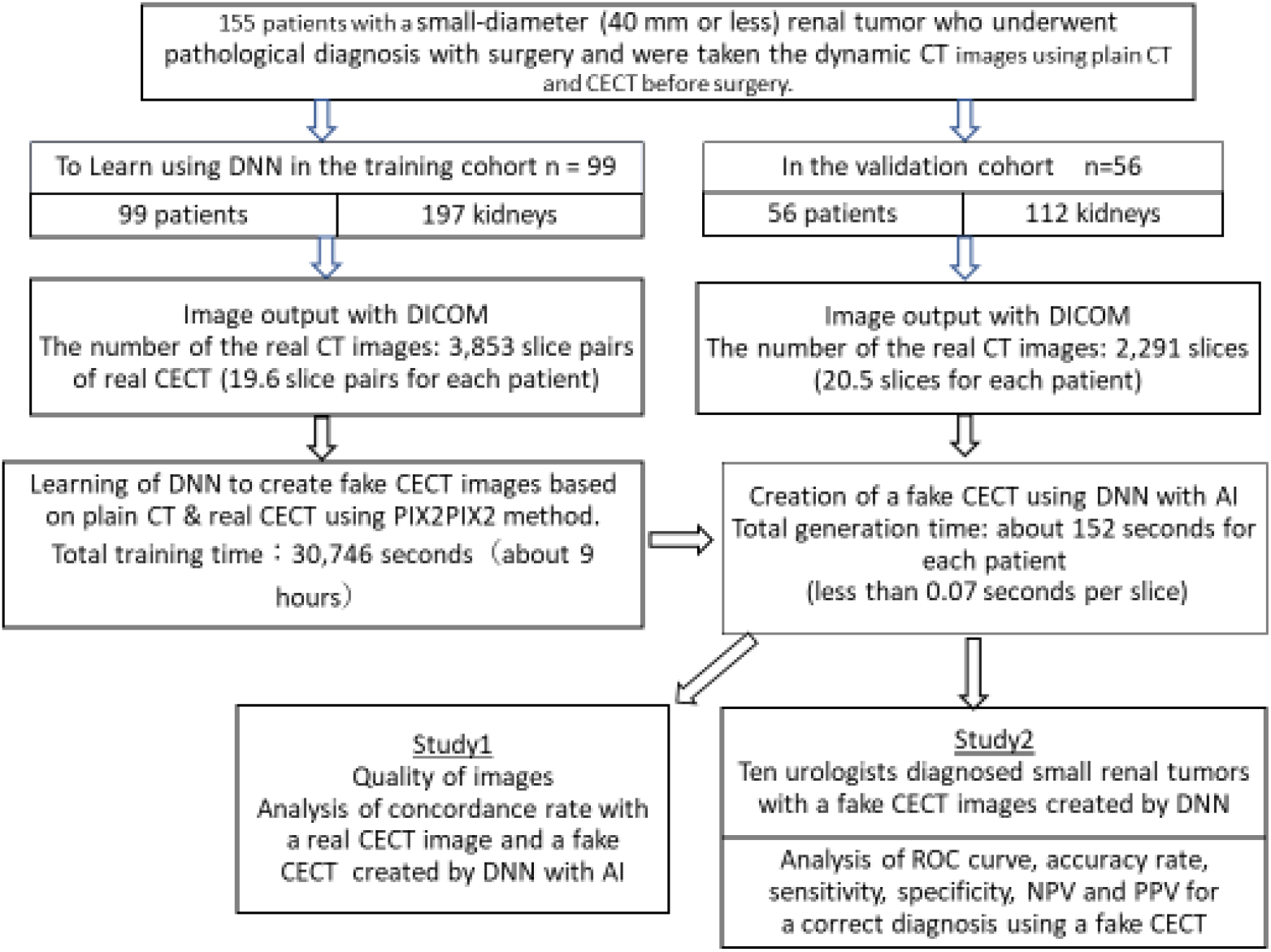
Flowchart of the study method. The left side shows the method in training cohort, and the right side shows the method in the validation cohort.

### Method to create a synthetic contrast-enhanced CT image using a learning DNN model

1. We used a group of plain CT and real CECT images (using only the arterial phase images) to create a DNN model that generated a synthetic CECT image. The plain CT and real CECT images were obtained in 5 mm and 1 mm increments, respectively.
2. Both image sets were labeled according to the presence or absence of tumors (Figure 2).
3. The position of the kidney varied between the CT and CECT images because of respiration. Therefore, we used the following method to reduce the variation in the position caused by the time lag when capturing kidney images:
  a. From the real CECT images, we selected an image that had z-coordinates near the z-coordinate of a plain CT image and that had the highest zero-mean normalized cross-correlation (ZNCC) with that plain CT image.

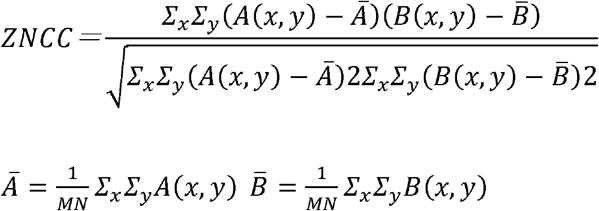 A (x, y): Luminance value at position (x, y) of image A B (x, y): Luminance value at position (x, y) of image B M: Number of pixels in the horizontal direction of images A and B N: Number of pixels in the vertical direction of images A and B
  b. We moved the selected real CECT image in the XY direction to acquire a higher concordance rate for the plain CT images. The shifted CECT image was then paired with a plain CT image.
  c. We created a training dataset by repeating steps (a) and (b) for the plain CT images of the group.
4. Using the above training data, we created a learned DNN using pix2pix, which is a popular method for image-to-image translation^21^.

**Figure 2.**
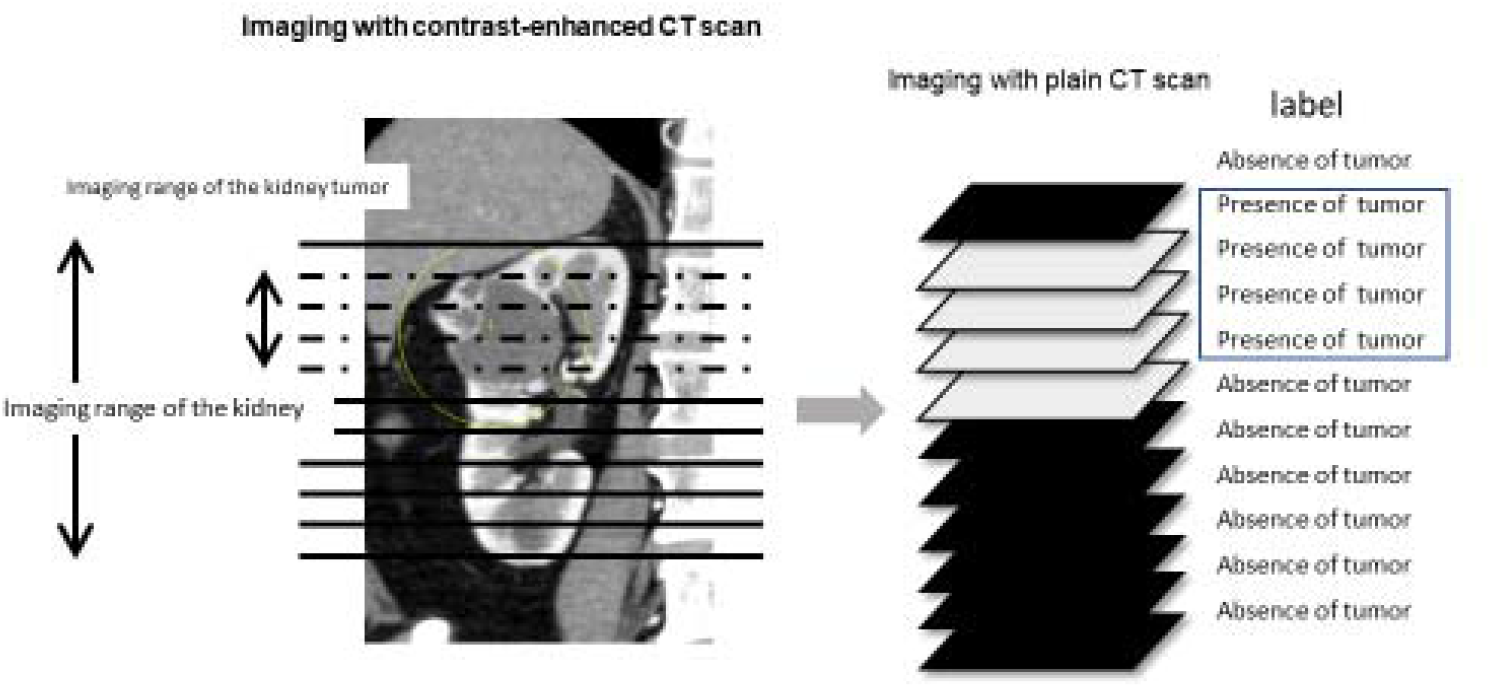
Plain and real contrast-enhanced computed tomography images for each computed tomography image labeled with the presence or absence of tumors.

Example of outputs from the learned DNN are shown in Figure 3 (in the figure: left, the real plain CT image; central, the synthetic CECT image created using the DNN; right, the real CECT image).

**Figure 3.**
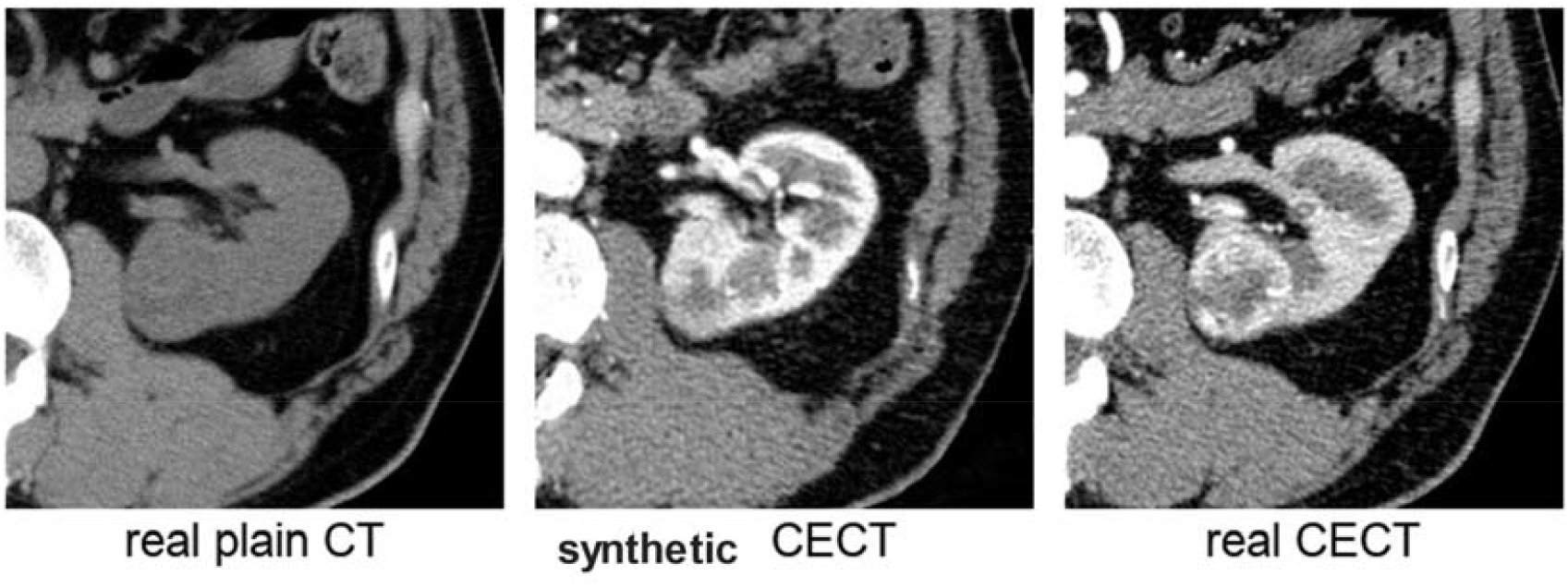
Example of the output of the learned deep neural network (DNN). (left-real plain computed tomography image, central-synthetic contrast-enhanced computed tomography [CECT] image created by DNN, and right-real CECT image)

### Methods of examination of the quality of synthetic CECT images created using learned DNN

#### Method of Study 1: analysis of concordance rate using ZNCC

1. Using the DNN, we created a synthetic CECT image from a plain CT image for evaluating against the real CECT image of each patient in the training cohort (n=56).
2. The concordance rate was evaluated using the ZNCC between the real and synthetic CECT images.
3. We decided on a central area and cut out 224 × 224 pixels of the synthetic CECT image, which originally had 256 × 256 pixels.
4. We moved the cropped-out area in the range of 16 pixels up, down, left, and right and determined the image location where the ZNCC value was the largest and recorded this value.
5. We analyzed the concordance rate between the real and synthetic CECT images using the above-mentioned equation for calculating ZNCC.

### Method of Study 2: diagnoses by urologists and receiver operating characteristic (ROC) curve analysis

We hypothesized that it was possible to diagnose small renal tumors using synthetic CECT images created by AI (Fig 1, right side).

The synthetic CECT images in the validation cohort dataset (n=56) with masked clinical information were evaluated by 10 urologists (with 20, 17, 14, 12, 12, 6, 5, 5, 4, and 3 years of clinical experience, respectively) individually. Ten urologists evaluated 2,291 CT images (approximately 20.5 images per patient) and indicated whether there were findings suggestive of a renal tumor using synthetic CECT and/or only plain CT.

### Statistical analysis

We analyzed the data using SPSS ® statistical software (ver20, Chicago, USA). The ROC curve, area under the curve (AUC), sensitivity, specificity, concordance rate, positive predictive value (PPV), and negative predictive value (NPV) were used for analysis. Statistical significance was set at *p*<0.05.

### Statistical analysis using ZNCC

ZNCC is the same as the Pearson’s correlation coefficient. Statistical analysis using ZNCC had shown that a valid image was created when the concordance rate was 70% or more.23,24

## Results

Using the DNN, we created synthetic CECT images for 99 patients (197 kidneys) in the training cohort and 56 patients (112 kidneys) in the validation cohort. The number of CT image slices and the training time for all kidneys were shown in Figure 1. The mean number of CT image slices for each kidney was 19.8 ± 3.0 sheets in the training cohort and 20.5 ± 2.4 sheets in the validation cohort. The number of image slices obtained for a renal tumor in each case was 4.7 ± 2.0 sheets in the training cohort (99 renal tumors) and 5.1 ± 1.8 sheets (56 renal tumors) in the validation cohort.

### Results of Study 1: examination of the quality of synthetic CECT images

Patient characteristics in the validation cohort are shown in Table 1. We compared the difference in ZNCC across patients with no renal tumor, renal tumor, exophytic renal tumor, and endophytic renal tumor. When compared with real CECT images, synthetic CECT images without a renal tumor, with a renal tumor, with an exophytic renal tumor, and with an endophytic renal tumor had mean ZNCCs of 0.767 ± 0.053, 0.770 ± 0.057, 0.779 ± 0.057, and 0.742 ± 0.062, respectively. Therefore, we created a suitable synthetic CECT image regardless of the presence of a renal tumor, exophytic renal tumor, or endophytic renal tumor.

**Table 1.**
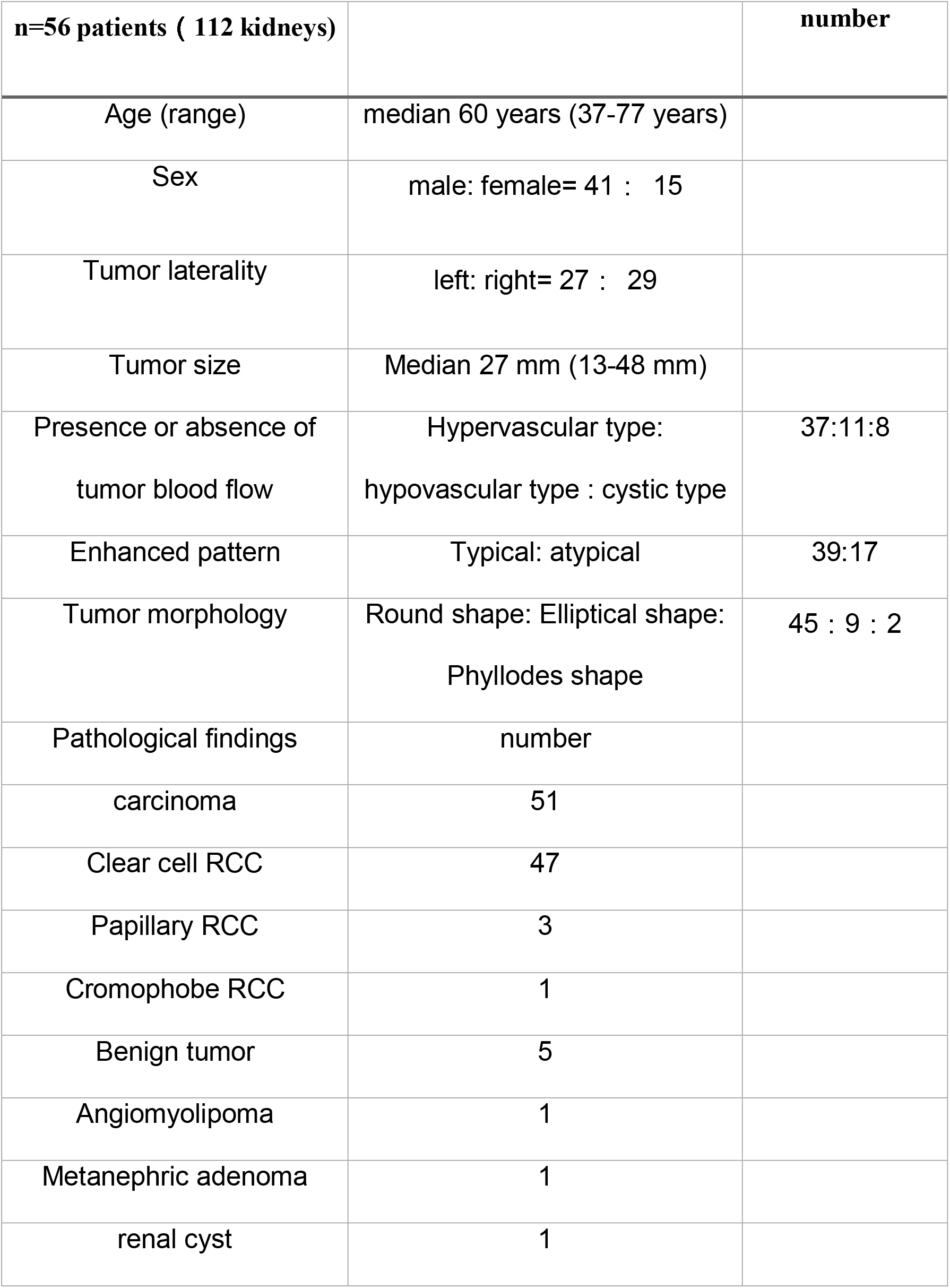
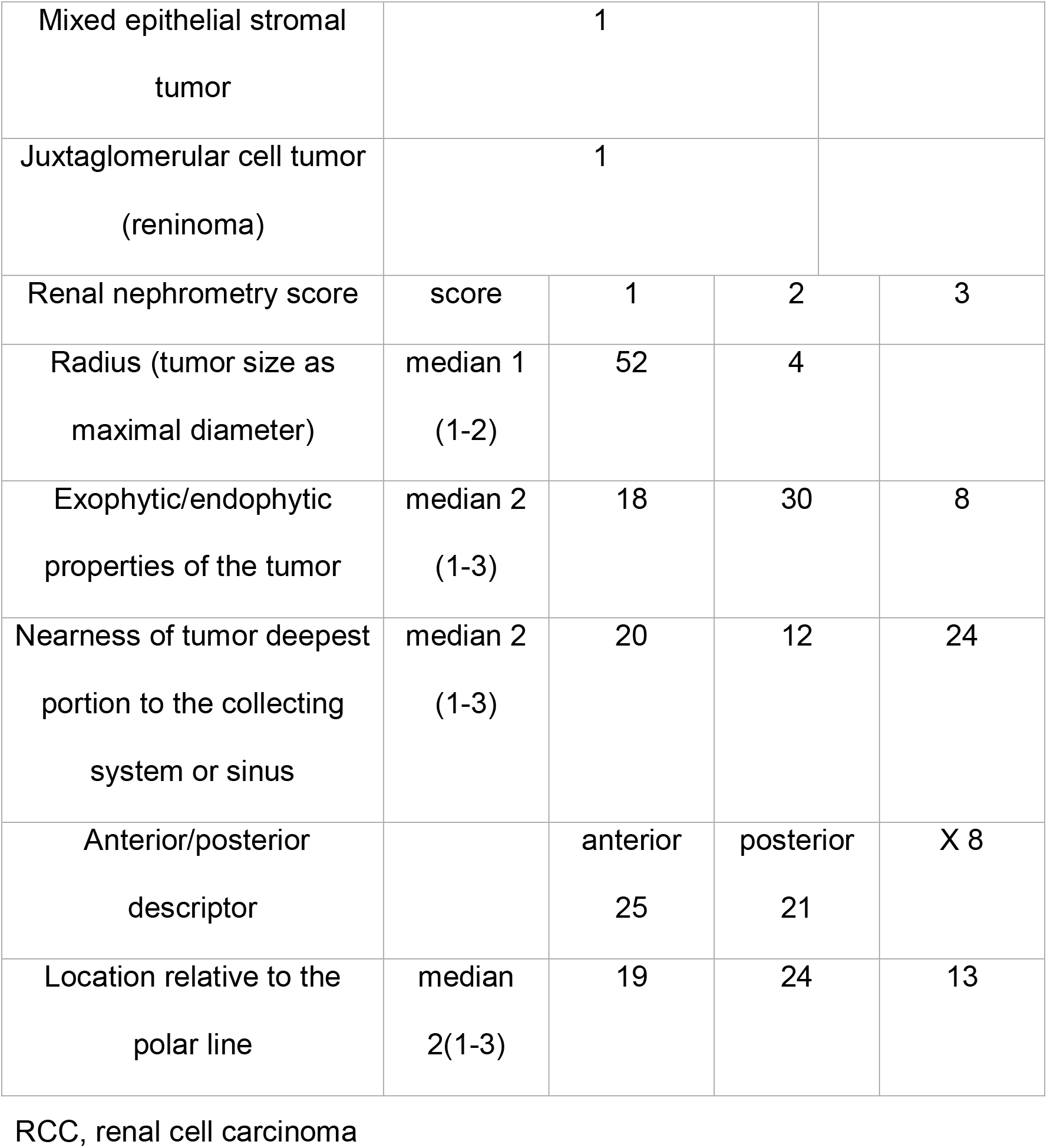
Patients characteristics

| **n=56 patients ( 112 kidneys)** |  | **number** |
| --- | --- | --- |
| Age (range) | median 60 years (37-77 years) |  |
| Sex | male: female= 41 : 15 |  |
| Tumor laterality | left: right= 27 : 29 |  |
| Tumor size | Median 27 mm (13-48 mm) |  |
| Presence or absence of<br>tumor blood flow | Hypervascular type:<br>hypovascular type : cystic type | 37:11:8 |
| Enhanced pattern | Typical: atypical | 39:17 |
| Tumor morphology | Round shape: Elliptical shape:<br>Phyllodes shape | 45 : 9 : 2 |
| Pathological findings | number |  |
| carcinoma | 51 |  |
| Clear cell RCC | 47 |  |
| Papillary RCC | 3 |  |
| Cromophobe RCC | 1 |  |
| Benign tumor | 5 |  |
| Angiomyolipoma | 1 |  |
| Metanephric adenoma | 1 |  |
| renal cyst | 1 |  |

|  |  |  |  |  |
| --- | --- | --- | --- | --- |
| Mixed epithelial stromal tumor |  | 1 |  |  |
| Juxtaglomerular cell tumor (reninoma) |  | 1 |  |  |
| Renal nephrometry score | score | 1 | 2 | 3 |
| Radius (tumor size as maximal diameter) | median 1 (1-2) | 52 | 4 |  |
| Exophytic/endophytic properties of the tumor | median 2 (1-3) | 18 | 30 | 8 |
| Nearness of tumor deepest portion to the collecting system or sinus | median 2 (1-3) | 20 | 12 | 24 |
| Anterior/posterior descriptor |  | anterior 25 | posterior 21 | X 8 |
| Location relative to the polar line | median 2(1-3) | 19 | 24 | 13 |

RCC, renal cell carcinoma

Subsequently, the difference between real and synthetic CECT images for each slice was calculated. Regarding the concordance rate for each image (n=2,291) (with tumor, without tumor, exophytic tumor, and endophytic tumor), slices without a tumor, with a tumor, with an exophytic tumor, and with an endophytic tumor had average ZNCCs of 0.773, 0.755, 0.766, and 0.717, respectively. The ZNCC exceeded 0.70 in all tumor morphologies, but the ZNCC with a synthetic CECT over slices with endophytic tumors tended to worsen the quality of the image.

### Results of Study 2: diagnosing small-diameter renal tumors using synthetic CECT images

We examined the judgments of 10 urologists and the concordance rates of the images created through AI. The accuracy, sensitivity, specificity, PPV, and NPV for an accurate diagnosis using a synthetic CECT image were 74.2%, 72.7%, 75.7%, 75.0%, and 73.5%, respectively. The concordance rate of six or more of the 10 evaluators diagnosing a renal tumor using a synthetic CECT image was 78.5% (44/56 kidneys). The concordance rate of six or more of the 10 urologists detecting no renal tumor was 83.9% (47/56 kidneys).

Considering the concordance rates for the accurate and inaccurate detection of a renal tumor by the urologists, the mean ZNCC of a synthetic CECT image that accurately predicted a renal tumor was 0.769 ± 0.066 when compared with a real CECT image. The mean ZNCC with a synthetic CECT image that inaccurately predicted a renal tumor was 0.766 ± 0.057 compared with a real CECT image. There was no synthetic CECT image with quality inferior to that of the real diagnostic image.

The ROC curve analysis for the concordance rate for the accurate answer revealed a high AUC of 0.892 (*p*<0.001) using synthetic CECT compared to that obtained with only plain CT (AUC=0.720, *p*<0.001). The quality of synthetic CECT images was sufficient for the urologists to distinguish between the presence and absence of a renal tumor.

## Comment

In this first report, we create a high-quality synthetic CECT image that was highly concordant with a real CECT image using DNN.

Abdominal ultrasonography is widely used as a screening method for renal cancer as RCCs are more frequently detected on abdominal ultrasonography than other solid cancer types. Additionally, the proportion of localized RCCs among accidentally diagnosed RCCs was 74.6% in one study, which was significantly higher than that among symptomatic RCCs (35.8%).^25^ However, abdominal ultrasonography alone is not sufficient to distinguish between renal angiomyolipoma and RCC; abdominal ultrasonography should be performed first as a screening method, followed by CT due to findings suggestive of renal cancer.^26^ CT is used as a definitive diagnostic method for RCC and has been found to be superior to abdominal ultrasonography, especially for visualizing small-diameter (≤ 3 cm) renal tumors.^27,28^

CECT is warranted for the definitive diagnosis of RCC. Urologists rely on the information obtained from CECT to formulate a treatment plan. MRI and positron emission tomography/CT are alternatives to CECT; however, their detection capabilities are inferior to those of CECT. We hypothesized that by creating a synthetic CECT image from a plain CT image, the problem of using a contrast medium can be solved while maintaining the image detection ability. We first verified the quality of synthetic CECT images created using deep-learned DNNs. The synthetic CECT image created using the DNN was appropriate when the concordance rate between the synthetic CECT image and the corresponding contrast-enhanced CT image was 70% or more. We found that the synthetic CECT images created using the DNN were similar to real CECT images, and a concordance rate of 70% or more could be obtained regardless of the presence or absence of a renal tumor. Thus, the synthetic CECT images were found to be sufficiently concordant for clinical use.

Furthermore, we investigated whether synthetic CECT images could be used for diagnosing renal tumors clinically. Assessments by 10 urologists revealed that synthetic CECT images created using DNN could be used for screening and diagnosis of RCC with sufficient accuracy, sensitivity, and specificity. This is a novel report in that it focuses on renal cancer screening using images created by image composition technology using DNN. To date, the purpose of imaging research using AI in the field of renal cancer has been to distinguish between benign and malignant kidney cancer,^15-18^ histological types of RCC, and nuclear grades of RCC.^19,20^ There have been no studies on the screening of small-diameter renal tumors using CT images created using DNN.

This study had several limitations. First, this was a retrospective study with a small sample size; the effectiveness of our image composition model should be confirmed in a larger number of cases. Second, our model was found to be useful for screening for RCC; however, it should be improved for use in the diagnosis and further treatment and surgical planning of benign/malignant renal tumors. Finally, as this study used images of small-diameter renal tumors, the potential of this model in detecting larger renal tumors remains unknown.

## Conclusions

This study is the first to create high-quality images from plain CT images using a DNN, which was generated through AI, with a high concordance rate on comparing with a real CECT image. The results suggest that synthetic CECT images can be used for urological diagnoses and clinical screening.

## Data Availability

All data produced in the present work are contained in the manuscript

## References

1. Finelli A, Ismaila N, Bro B, et al. Management of small renal masses: American Society of Clinical Oncology clinical practice guideline. J Clin Oncol. 2017;35(6):668–680.

2. Novara G, Ficarra V, Antonelli A, et al. Validation of the 2009 TNM version in a large multi-institutional cohort of patients treated for renal cell carcinoma: Are further improvements needed? Eur Urol. 2010;58(4):588–595.

3. Jayson M, Sanders H. Increased incidence of serendipitously discovered renal cell carcinoma. Urology. 1998;51(2):203–205.

4. Chu JS, Wang ZJ. Protocol optimization for renal mass detection and characterization. Radiol Clin North Am. 2020;58(5):851–873.

5. Sasaguri K, Takahashi N. CT and MR imaging for solid renal mass characterization. Eur J Radiol. 2018;99:40–54.

6. Israel GM, Bosniak MA. Pitfalls in renal mass evaluation and how to avoid them. RadioGraphics. 2008;28(5):1325–1338.

7. Shao P, Tang L, Li P, et al. Precise segmental renal artery clamping under the guidance of dual-source computed tomography angiography during laparoscopic partial nephrectomy. Eur Urol. 2012;62(6):1001–1008.

8. Janus CL, Mendelson DS. Comparison of MRI and CT for study of renal and perirenal masses. Crit Rev Diagn Imaging. 1991;32(2):69–118.

9. Dallmer JR, Robles J, Wile GE, Koyama T, Barocas DA. The harms of hematuria evaluation: Modeling the risk-benefit of using split bolus computerized tomography urography to reduce radiation exposure in a theoretical cohort. J Urol. 2019;202(5):899–904.

10. Capogrosso P, Capitanio U, La Croce G, et al. Follow-up after treatment for renal cell carcinoma: The evidence beyond the guidelines. Eur Urol Focus. 2016;1(3):272–281.

11. Neisius A, Wang AJ, Wang C, et al. Radiation exposure in urology: A genitourinary catalogue for diagnostic imaging. J Urol. 2013;190(6):2117–2123.

12. Lipsky MJ, Shapiro EY, Hruby GW, McKiernan JM. Diagnostic radiation exposure during surveillance in patients with pT1a renal cell carcinoma. Urology. 2013;81(6):1190–1195.

13. Dilauro M, Quon M, McInnes MD, et al. Comparison of contrast-enhanced multiphase renal protocol CT versus MRI for diagnosis of papillary renal cell carcinoma. AJR Am J Roentgenol. 2016;206(2):319–325.

14. Ljungberg B, Albiges L, Abu-Ghanem Y, et al. European Association of Urology guidelines on renal cell carcinoma: The 2019 update. Eur Urol. 2019;75(5):799–810.

15. Oberai A, Varghese B, Cen S, et al. Deep learning based classification of solid lipid-poor contrast enhancing renal masses using contrast enhanced CT. Br J Radiol. 2020;93(1111):20200002.

16. Baghdadi A, Aldhaam NA, Elsayed AS, et al. Automated differentiation of benign renal oncocytoma and chromophobe renal cell carcinoma on computed tomography using deep learning. BJU Int. 2020;125(4):553–560.

17. Kocak B, Kaya OK, Erdim C, Kus EA, Kilickesmez O. Artificial intelligence in renal mass characterization: A systematic review of methodologic items related to modeling, performance evaluation, clinical utility, and transparency. AJR Am J Roentgenol. 2020;215(5):1113–1122.

18. Han S, Hwang SI, Lee HJ. The classification of renal cancer in 3-phase CT images using a deep learning method. J Digit Imaging. 2019;32(4):638–643.

19. Kocak B, Durmaz ES, Ates E, Kaya OK, Kilickesmez O. Unenhanced CT texture analysis of clear cell renal cell carcinomas: A machine learning-based study for predicting histopathologic nuclear grade. AJR. 2019:W1–W8.

20. Shu J, Wen D, Xi Y, et al. Clear cell renal cell carcinoma: Machine learning-based computed tomography radiomics analysis for the prediction of WHO/ISUP grade. Eur J Radiol. 2019;121:108738.

21. Kaji S, Kida S. Overview of image-to-image translation by use of deep neural networks: Denoising, super-resolution, modality conversion, and reconstruction in medical imaging. Radiol Phys Technol. 2019;12(3):235–248.

22. Kutikov A, Uzzo RG. The RENAL nephrometry score: A comprehensive standardized system for quantitating renal tumor size, location and depth. J Urol. 2009;182(3):844–853.

23. Mukaka MM. Statistics corner: A guide to appropriate use of correlation coefficient in medical research. Malawi Med J. 2012;24(3):69–71.

24. Chen H. YJaJZ. 3D Reconstruction for Robot Navigation Based on Projection of Virtual Height Line and Its Performance Evaluation International Conference on Measuring Technology and Mechatronics Automation; vol 1; 2010:3–6, 2020.

25. Lightfoot N, Conlon M, Kreiger N, et al. Impact of noninvasive imaging on increased incidental detection of renal cell carcinoma. Eur Urol. 2000;37(5):521–527.

26. Einstein DM, Herts BR, Weaver R, Obuchowski N, Zepp R, Singer A. Evaluation of renal masses detected by excretory urography: Cost-effectiveness of sonography versus CT. AJR Am J Roentgenol. 1995;164(2):371–375.

27. Curry NS. Imaging the small solid renal mass. Abdom Imaging. 2002;27(6):629–636.

28. Jamis-Dow CA, Choyke PL, Jennings SB, Linehan WM, Thakore KN, Walther MM. Small (< or = 3-cm) renal masses: detection with CT versus US and pathologic correlation,” Radiology,. 1996;198(3):785–788.

